# Clinically Meaningful Cognitive Improvement from Intermittent Neurostimulation of the Nucleus Basalis of Meynert in Parkinson’s Disease

**DOI:** 10.64898/2026.11.02.26366824

**Authors:** Kevin B. Wilkins, Prerana Acharyya, Charlotte Casselton, Annie K. Abay, Rachel A. Crockett, Gayle K. Deutsch, Mahsa Malekmohammadi, Hemant Bokil, Vincent Cina, Brandon Reid, Ashwin G. Ramayya, Michael Zeineh, Gang Seo, Tatianna M. Howard, Maria Papworth Burrel, David T. Blake, Jaimie M. Henderson, Vivek P. Buch, Helen Bronte-Stewart

## Abstract

Cognitive decline begins in early stages of Parkinson’s disease (PD) for many individuals and remains largely untreated. The nucleus basalis of Meynert (NBM), the brain’s cortical cholinergic hub, and its lateral efferent fiber bundle degenerate early in PD and are linked to cognitive and cognitive-motor impairment. Pre-clinical evidence indicates intermittent stimulation of the NBM improves cognition. The objective of this study is to establish the feasibility of combined continuous subthalamic nucleus (STN) and intermittent NBM DBS for cognitive impairment in people with PD. Five patients with PD and cognitive impairment approved for STN DBS were enrolled as part of an ongoing clinical trial for combined STN and NBM DBS under an Investigational Device Exemption. Three participants received a ‘vertical’ trajectory in which two leads were placed in the STN, and two additional leads were placed in the anterior portion of the NBM, both using the same burr hole for each hemisphere. The two other participants received a novel ‘lateral trajectory’ in which the NBM leads were placed along the NBM lateral efferent bundle via a small twist craniostomy behind the ear. An automated programming algorithm DBS Illumina 3D™ (Boston Scientific) was used to program the NBM leads based on volume of tissue activated modeling. The investigational Chronos software (Boston Scientific) was used to deliver continuous high frequency DBS to the STN 24 hours/day and intermittent patterned DBS to the NBM for 1 hour/day. The two leads placed in the NBM (‘Vertical’ target) or its lateral efferent bundle (‘Lateral’ target) were successfully and accurately placed as part of the usual STN DBS procedure. All participants were successfully programmed using the Illumina 3D™ algorithm without adverse effects. Following 6 months of combined continuous STN and intermittent NBM DBS, all five participants saw clinically meaningful improvements in cognition as part of a neuropsychological assessment battery, as well as the expected motoric benefit from STN DBS. The current study demonstrated that intermittent stimulation of the NBM to target cortical cholinergic deficits may provide clinically meaningful improvements in cognitive symptoms in Parkinson’s disease. ClinicalTrials.gov identifier: NCT05968703

## Introduction

Cognitive decline begins in early stages of Parkinson’s disease (PD) and progresses to dementia in approximately 50% of people with PD after ten years and over 80% after 20 years^1–4^. Dopaminergic medication and deep brain stimulation (DBS) provide long-term improvement of cardinal motor symptoms in PD, but cognition remains largely unaddressed despite a long window for potential intervention before dementia occurs^5^. The only currently approved treatment for worsening cognition in PD are cholinesterase inhibitors, which are of limited efficacy and can cause troublesome side effects^6^. Treatment for cognitive impairment typically only starts when it reaches later stages of mild cognitive impairment or dementia, by which time significant cortical cholinergic denervation results in marginal presence of acetylcholine remaining in cortical synaptic clefts, thus limiting the effect of any acetylcholine-enhancing therapy^1^. There is currently no approved treatment that reverses or even stabilizes cognitive decline in PD.

The cortical cholinergic network plays a crucial role in cognitive domains such as attention, visuospatial ability, and executive function^7^. The nucleus basalis of Meynert (NBM) is considered the cortical cholinergic hub as it is the primary source of cholinergic innervation to the cortex^8^. The NBM is comprised of approximately 90% cholinergic neurons and has two major outflow fiber tracts, the medial and lateral efferent fiber bundles, which provide cholinergic innervation to vast expanses of the cortical mantle, including the parolfactory, cingulate, pericingulate, and retrosplenial cortices, as well as the frontoparietal operculum, insula, superior temporal gyrus, and other portions of the frontal, parietal, and temporal cortex^9,10^. Post-mortem studies in PD show significant loss of cholinergic neurons in the NBM and degeneration of its cholinergic fiber projections^11,12^. The NBM is known to degenerate early in PD, with 32% cell loss in non-demented patients, rising to 54-70% in PD dementia^12–18^. In-vivo MRI and PET imaging have found similar results and shown that degeneration of the NBM and its fiber projections is predictive of cognitive impairment^19–24^. This degeneration appears to precede development of substantial cognitive symptoms^20^. Similarly, cognitive-motor impairments such as non-dopaminergic aspects of gait are also related to NBM degeneration^25,26^.

The NBM has been identified as a potential therapeutic neurostimulation target for cognitive-related symptoms^27,28^. A substantial body of pre-clinical literature over the last thirty-five plus years has demonstrated that neurostimulation of the NBM increases cortical acetylcholine and neurotrophic growth factor release and increases cerebral blood flow in rodents^29–33^. NBM stimulation in non-human primates has demonstrated improvements in working and spatial memory and sustained attention on the order of 30-45% improvement, which has been replicated in rodent models of cognitive impairment^34–41^. Crucially, these beneficial effects were only seen with intermittent stimulation (60 Hz pulse train for 10-20 seconds followed by 40-50 seconds of no stimulation for only 1 hour/day) and performance was impaired during continuous stimulation^34^. Intermittent stimulation is hypothesized to efficiently maximize cortical acetylcholine release and promote longer neuroplastic effects, whereas continuous stimulation has been shown to deplete neurotransmitter reserves^40,42^. This highlights a very different approach to neuromodulation of the cortical cholinergic network to treat the cognitive-motor syndrome compared to traditional DBS approaches to treat motor symptoms in PD.

Despite the positive therapeutic potential of NBM DBS for cognition in pre-clinical studies, the clinical studies in PD have had mixed efficacy^43–48^, which may be due to three major limitations: 1) A potentially “too little too late” timing of NBM DBS, when people with PD already had dementia and thus likely substantial cholinergic cell loss; 2) A single vertical trajectory to target both motor globus pallidus interna (GPi) and a portion of the NBM that may have resulted a suboptimal volume of tissue activated (VTA)^49^; and 3) The use of continuous neurostimulation, which impaired cognition in preclinical studies, instead of intermittent neurostimulation (60 Hz, cycling 10 sec on, 40 sec off for only 1 hour/day) that in preclinical studies has demonstrated increased cortical blood flow, cortical cholinergic tone, release of growth factors, and best performance. We sought to apply the three predictors of a successful outcome for motor (dopaminergic) DBS to cognitive (cholinergic) DBS: 1) select well-characterized candidates before the stage of dementia; 2) optimize target selection, lead location, and the generated VTA; and 3) use intermittent neurostimulation patterns which are optimized for the cognitive symptoms.

Here we present the initial proof of concept for the feasibility of optimizing surgical trajectories and stimulation parameters for DBS of the NBM for cognitive impairment in PD and the resulting preliminary efficacy. We designed and implemented two separate trajectories for the NBM, one based on the pre-clinical non-human primate evidence and one optimized for the unique geometry of the NBM and its lateral efferent bundle in the human brain^50^. An imaging-based pipeline and automated optimization algorithm was then utilized to determine optimal DBS settings for overlapping with the target NBM structure and minimizing overlap with nearby regions that could elicit side effects based on subject-specific lead location and anatomy. Neuropsychological outcomes were evaluated after six months of combined STN and intermittent NBM DBS.

## Results

Five participants approved for STN DBS and showing deficits in at least one test from comprehensive neuropsychological testing done as part of the standard Stanford DBS evaluation pipeline were enrolled in the study. Information on participant demographics is shown in Table 1.

**Table 1.**
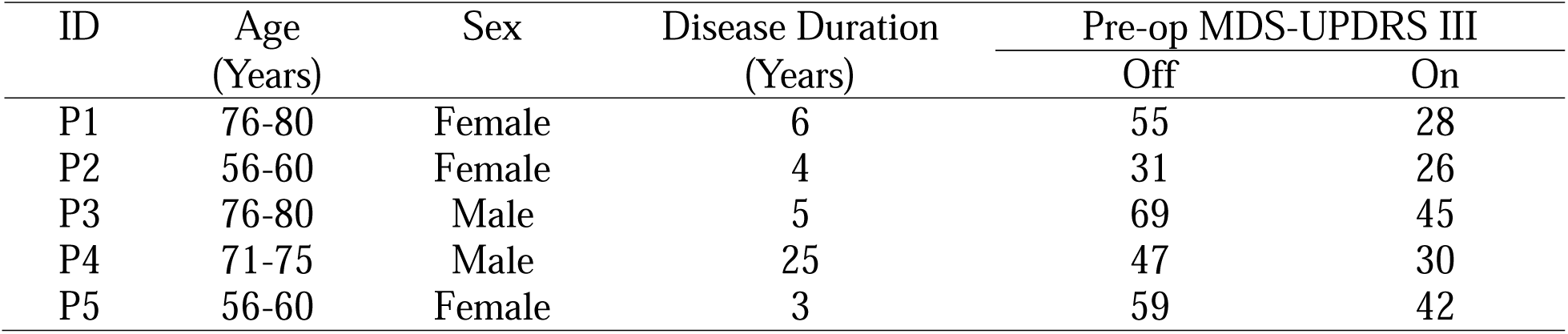
Participant Demographics.

| ID | Age<br>(Years) | Sex | Disease Duration<br>(Years) | Pre-op MDS-UPDRS III |  |
|---|---|---|---|---|---|
|  |  |  |  | Off | On |
| P1 | 76-80 | Female | 6 | 55 | 28 |
| P2 | 56-60 | Female | 4 | 31 | 26 |
| P3 | 76-80 | Male | 5 | 69 | 45 |
| P4 | 71-75 | Male | 25 | 47 | 30 |
| P5 | 56-60 | Female | 3 | 59 | 42 |

### Study Design

The current study is part of an ongoing single-site open label pilot clinical trial testing the safety and tolerability of the combination of continuous STN DBS and intermittent NBM DBS for cognitive and cognitive-motor deficits in people with PD (NCT05968703). Patients were randomized to either receive a Vertical Trajectory targeting the anterior portion of the NBM or a Lateral Trajectory targeting the NBM lateral efferent fiber bundle. All participants were implanted with multinodal DBS in the STN and NBM using four leads in the same procedure (Figure 1)^51^. Baseline assessments occurred approximately 1 month after surgery, prior to Initial Programming, with a follow-up assessment after 6 months of combined STN and NBM DBS.

**Figure 1.**
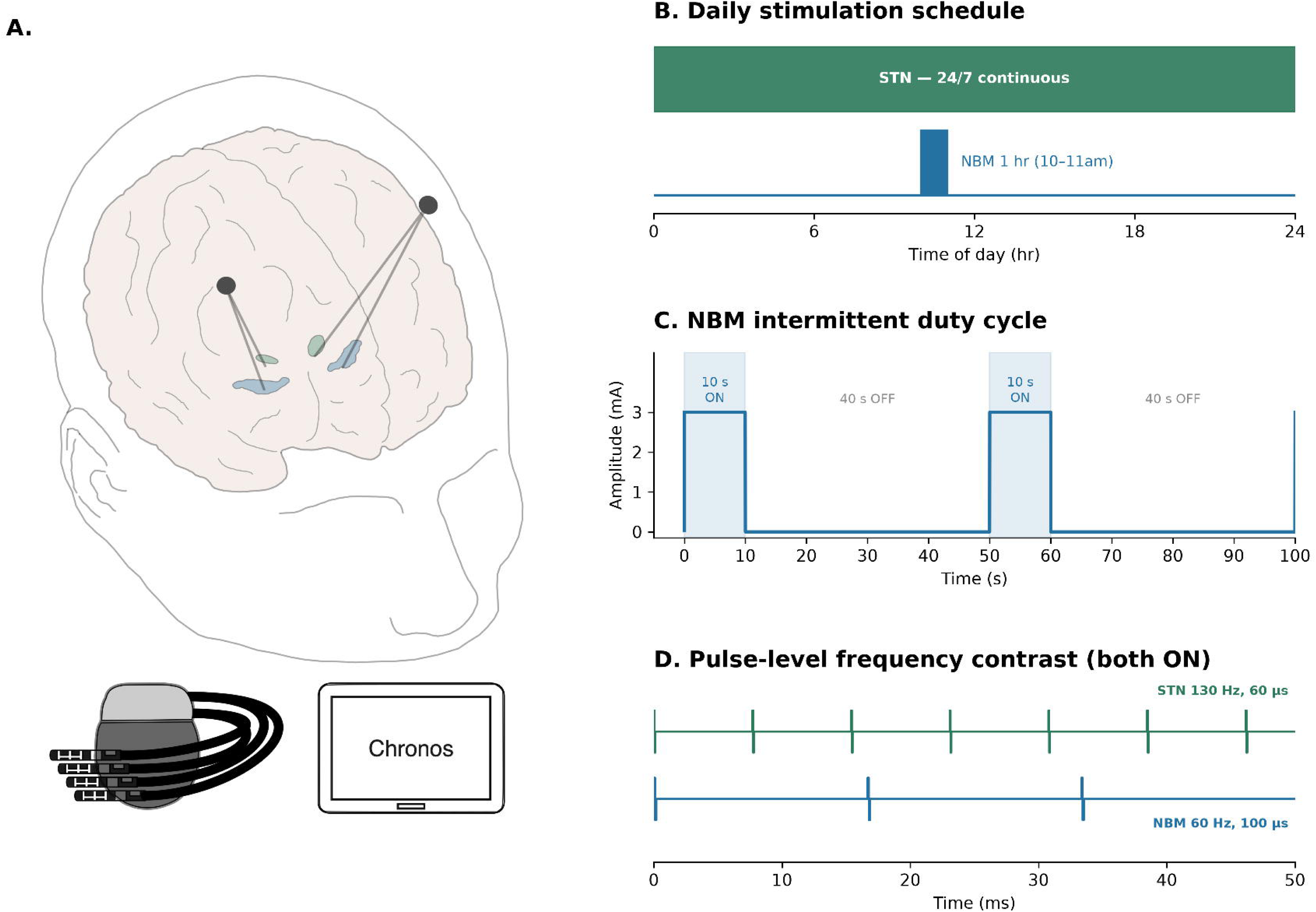
Overview of DBS Setup. (A) Illustration of the four DBS lead setup with the STN in green and NBM in blue using the vertical trajectory where the STN and NBM leads use the same burr hole. All four leads are connected to one 32 channel IPG controlled by the investigational Chronos software. (B) Schematic of the daily stimulation schedule. (C) Depiction of the intermittent cycling NBM DBS stimulation cycling 10 sec ON and 40 sec OFF during the 1 hour of active NBM stimulation. (D) Zoomed in trace showing the difference in frequency and pulse width between the STN (130 Hz, 60 μs) and NBM (60 Hz, 100 μs) leads.

### Designing Surgical Trajectories

A vertical trajectory was designed to target the anterior portion of the NBM. The placement of the vertical NBM leads used the same burr hole as was used to place the STN leads. This trajectory was modeled after previous literature in the non-human primate that demonstrated consistent cognitive improvement when targeting 2 mm posterior to the crossing of the globus pallidus by the anterior commissure, and in the floor of the Globus Pallidus at the intersection of the Globus Pallidus Internus (GPi) and Externus (GPe)^34,35,37,52^. The current target followed suit, while considering human histological evidence for the highest density of cholinergic neurons approximately 4 mm posterior to the anterior commissure^9^. This target is approximately a 5 mm distance from the standard clinical target for GPi.

Vertical NBM trajectories have been used in previous approaches but may not be optimal for targeting the NBM. Considering the NBM is a horizontal, open, nearly flat area of the basal forebrain adjacent to and below the floor of the globus pallidus, the vertical approach places only one level of electrodes on a DBS directional lead within the target and limits the VTA to a small volume of the NBM. However, the diffuse NBM channels much of its outflow into a tight lateral efferent fiber bundle, which projects to the dorsal frontoparietal neocortex, middle and inferior gyri, inferotemporal cortex, parahippocampal gyrus, frontoparietal operculum, insula, and superior temporal gyrus via the external capsule, claustrum, and uncinate fasciculus^10^. Moreover, this bundle is located on the lateral side of the NBM and is thus a surgically approachable target. Therefore, a lateral trajectory that aligns the axis of the DBS lead with the NBM outflow to the lateral efferent bundle may allow the VTA to encompass most of the efferent fiber bundle, thus maximizing widespread cortical acetylcholine release.

### Successful DBS Lead Placement Across Both Trajectories

Patients underwent a modified two stage DBS implantation procedure with all 4 leads placed in stage one, followed by a 32-channel rechargeable implantable pulse generator (IPG) in stage two. Three patients received the Vertical Trajectory and two received the Lateral Trajectory. For the Lateral trajectory, the NBM lateral efferent fiber bundle was reconstructed from a diffusion weighted imaging scan using a previously validated individualized tractography pipeline^50^. The lateral trajectory was approached via a small twist-drill craniostomy immediately behind the ear, with an anteromedial trajectory to the lateral efferent bundle target. Intraoperative imaging (Loop-X®, Brainlab) was performed on a 15mm above cannula to confirm that the planned trajectory resulted in sufficient distance from the middle cerebral artery before the final placement of the lead at target depth. Importantly, a 0mm above or “to target” cannula was not used, to prevent a rigid object traversing past the deep Sylvian cistern.

All five patients successfully underwent the DBS procedure with lead location confirmed both immediately post-op and on a follow-up CT one month after surgery. Figure 2 shows placement of the DBS leads within the STN and within the anterior portion of the NBM on both sides for the Vertical Trajectory in Participant 1. Figure 3 shows placement of bilateral STN and NBM DBS leads, where the NBM leads were aligned with the intersection between the NBM and its immediate lateral efferent fiber bundle outflow using the Lateral Trajectory in Participant 2. MNI coordinates for the center of each of the four electrode levels for all participants are shown in Table 2.

**Figure 2.**
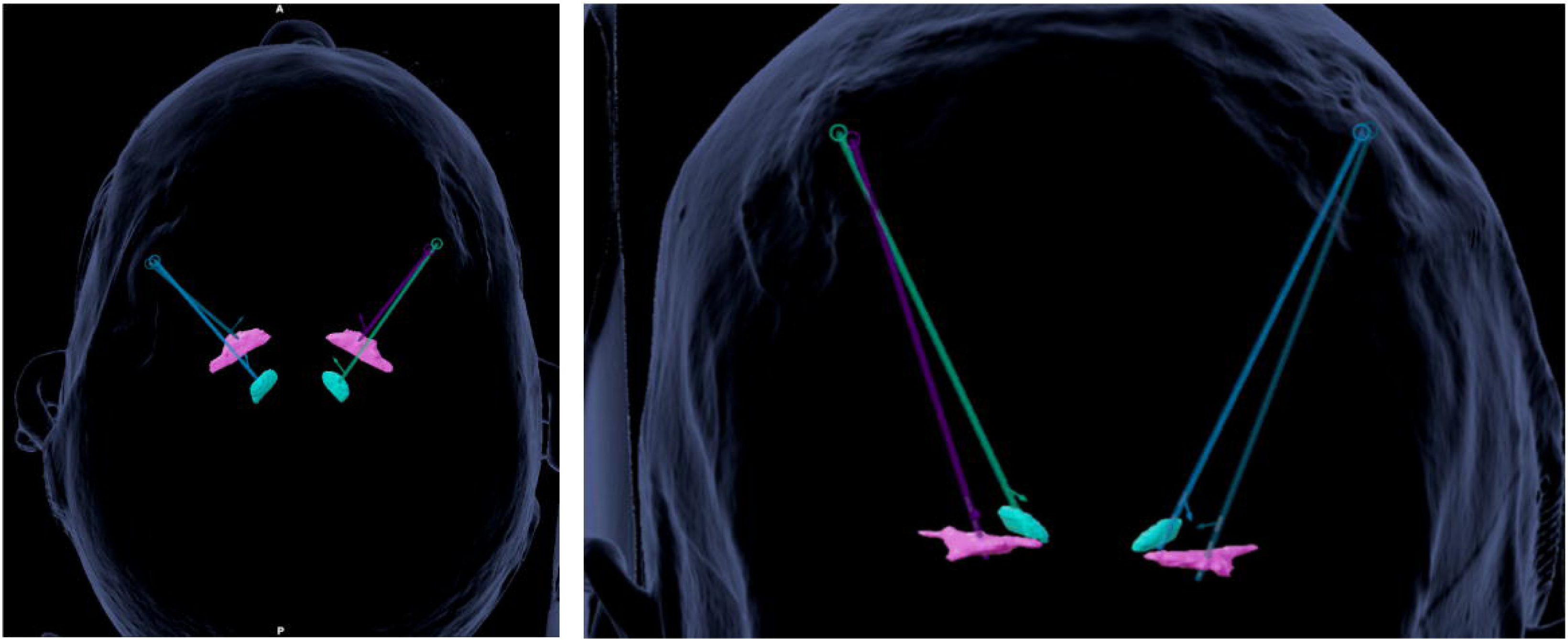
Lead reconstructions for the Vertical Trajectory. (Left) Axial and (Right) coronal view of the four DBS leads targeting the STN (cyan) and NBM (magenta).

**Figure 3.**
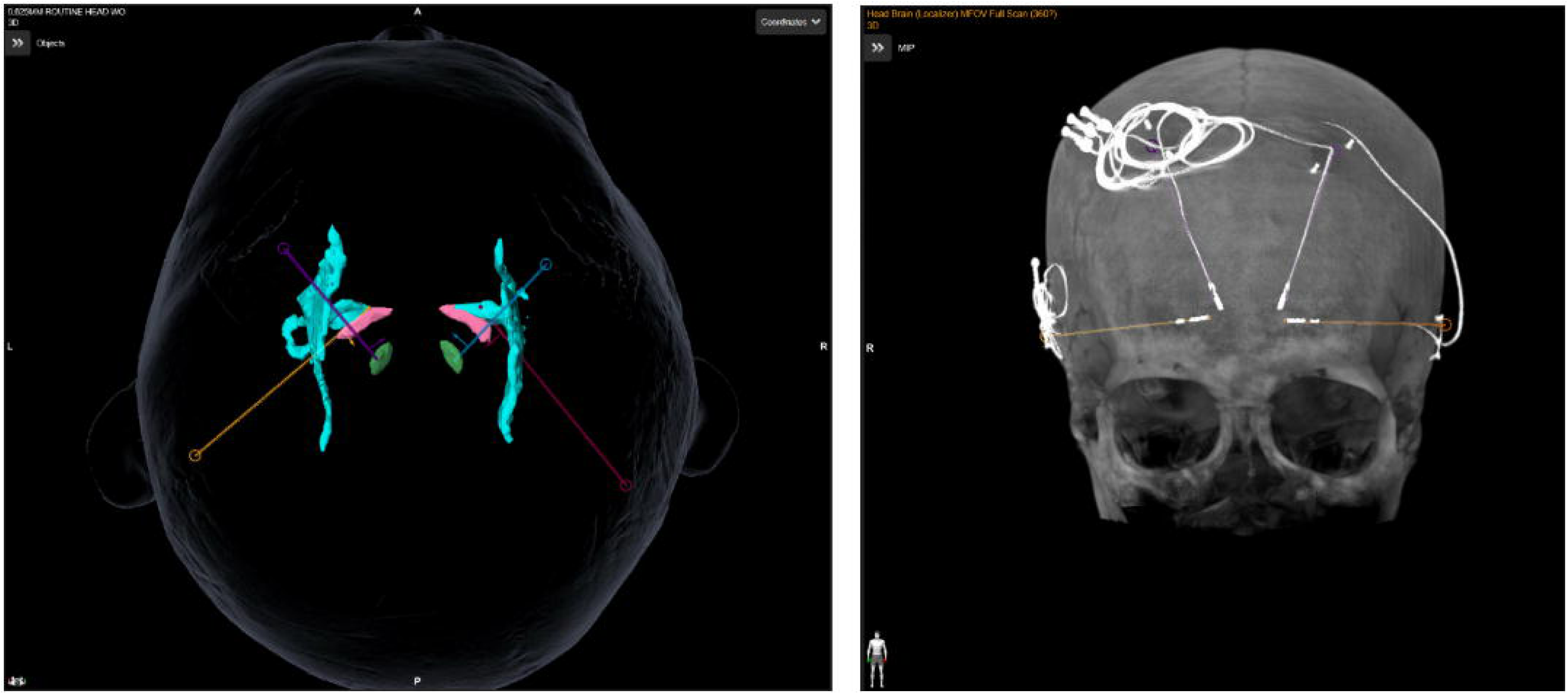
Lead reconstructions for the Lateral Trajectory. (Left) Axial view of the four DBS leads targeting the STN (green) and NBM (pink) and its lateral tract (cyan). (Right) Maximum intensity projection image from the Loop-X following the DBS procedure showing the four DBS leads.

**Table 2.** NBM MNI Coordinates.

| Vertical Trajectory |  |  |  |  |  |  |  |
|---|---|---|---|---|---|---|---|
|  |  | X |  | Y |  | Z |  |
|  |  | Left | Right | Left | Right | Left | Right |
| P1 | 1<sup>st</sup> Row | -18.3 | 17.8 | 11.8 | 11.1 | -9.0 | -5.3 |
|  | 2<sup>nd</sup> Row | -18.9 | 18.3 | 12.4 | 11.8 | -7.1 | -3.5 |
|  | 3<sup>rd</sup> Row | -19.6 | 18.9 | 12.9 | 12.5 | -5.3 | -1.7 |
|  | 4<sup>th</sup> Row | -20.2 | 19.4 | 13.5 | 13.1 | -3.5 | -0.1 |
| P4 | 1<sup>st</sup> Row | -21.3 | 25.0 | 6.2 | 5.6 | -9.8 | -8.8 |
|  | 2<sup>nd</sup> Row | -21.8 | 25.5 | 7.0 | 6.4 | -7.1 | -7.1 |
|  | 3<sup>rd</sup> Row | -22.3 | 26.0 | 7.8 | 7.3 | -5.4 | -5.4 |
|  | 4<sup>th</sup> Row | -22.9 | 26.5 | 8.6 | 8.2 | -3.7 | -3.6 |
| P5 | 1<sup>st</sup> Row | -18.9 | 17.0 | 9.4 | 9.9 | -9.1 | -8.4 |
|  | 2<sup>nd</sup> Row | -19.9 | 17.6 | 10.1 | 10.6 | -7.4 | -6.8 |
|  | 3<sup>rd</sup> Row | -19.7 | 18.4 | 10.8 | 11.3 | -5.5 | -5.0 |
|  | 4<sup>th</sup> Row | -20.1 | 19.3 | 11.5 | 11.9 | -3.8 | -3.3 |
| Lateral Trajectory |  |  |  |  |  |  |  |
| P2 | 1<sup>st</sup> Row | -13.2 | 14.7 | 10.0 | 10.8 | -6.0 | -4.8 |
|  | 2<sup>nd</sup> Row | -15.1 | 16.5 | 9.3 | 10.2 | -6.3 | -5.2 |
|  | 3<sup>rd</sup> Row | -17.0 | 18.4 | 8.7 | 9.6 | -6.5 | -5.5 |
|  | 4<sup>th</sup> Row | -18.8 | 20.2 | 8.0 | 9.0 | -6.8 | -5.9 |
| P3 | 1<sup>st</sup> Row | -16.0 | 20.5 | 13.6 | 13.9 | -5.1 | -5.7 |
|  | 2<sup>nd</sup> Row | -17.5 | 21.7 | 12.4 | 12.5 | -5.5 | -6.0 |
|  | 3<sup>rd</sup> Row | -19.0 | 22.9 | 11.1 | 10.9 | -5.9 | -6.3 |
|  | 4<sup>th</sup> Row | -20.5 | 24.2 | 9.8 | 9.4 | -6.2 | -6.7 |

\*Shown as the stereotactic coordinates (mm) with reference to the midcommissural point of the anterior-commissure-posterior commissure plane

### Image-Guided Stimulation Parameters

An automated modeling- and imaging-based approach was used to determine the NBM stimulation configuration since, unlike STN DBS for motor symptoms, acute stimulation-related behavior changes cannot be used to guide setting up NBM DBS. Therefore, the DBS Illumina 3D™ algorithm (Boston Scientific) was used^53^. Illumina 3D™ is an optimization algorithm that uses patient-specific imaging to find the stimulation settings that maximizes overlap with a target region of interest (ROI) while minimizing stimulation of regions of avoidance (ROAs) that could result in unknown or undesired side effects, all within stimulation charge density, total charge, and amplitude safety limits. The algorithm allows user-selected weightings that set the priority of not stimulating the avoidance regions (controlled by a slider to set ‘avoidance ratio’), as well as the priority of reduced Stimulation Field Model (SFM) volume (controlled by a slider to set the ‘background ratio’).

The Optic Tract, Amygdala, and Globus Pallidus Externus (GPe) were set as ROAs for both the Vertical and Lateral Trajectory, whereas the NBM was set as the ROI for the Vertical Trajectory and one combined object consisting of the NBM and the reconstructed lateral NBM tract was set as the ROI for the Lateral Trajectory. The Illumina 3D™ algorithm was run across 11 different weightings for both the avoidance ratio and background ratio, resulting in a total of 121 different potential stimulation configurations. For each configuration, the percent overlap with the ROI and each ROA was determined, as well as the overall spill (i.e., portion of the SFM that does not overlap with the ROI). Stimulation configurations with excessive overlap of ROAs were not considered. Visualizations were created of the remaining potential configurations comparing the ROI overlap compared to overall spill volume to identify the best candidates that efficiently overlapped with the target. Visualizations of the resulting SFMs for the candidate configurations alongside the nearby ROAs were then produced.

Figure 4 shows the ROI overlap compared to Spill Volume for all the potential Illumina-recommended stimulation settings across different avoidance and background ratios in Participant 1. Settings that excessively overlapped with any of the ROAs are depicted with ‘X’s and were not considered further. In cases where potential stimulation settings offered similar ROI overlap, the setting that minimized Spill volume was prioritized. The red box indicates the clinician-determined optimal selected setting that maximized ROI overlap, while minimizing spill. The subsequent stimulation configuration and resulting SFM for each of the left and right NBM are shown in Figure 4. The SFM was largely centered on the second level with minimal directional steering given the central placement within the target for both Vertical trajectories. Meanwhile, the Lateral trajectory in Participant 2 utilized directional electrode activation to steer the electric field so that it optimally included the NBM lateral outflow and fiber bundle (Figure 5). The location of the lead relative to nearby ROAs allowed a higher overall stimulation current (3.1/3.4 mA for left/right) and ROI overlap (75/100 mm^3^) without excessive overlap of any ROAs compared to the Vertical trajectory (2.3/2.6 mA and 31/51 mm^3^).

**Figure 4.**
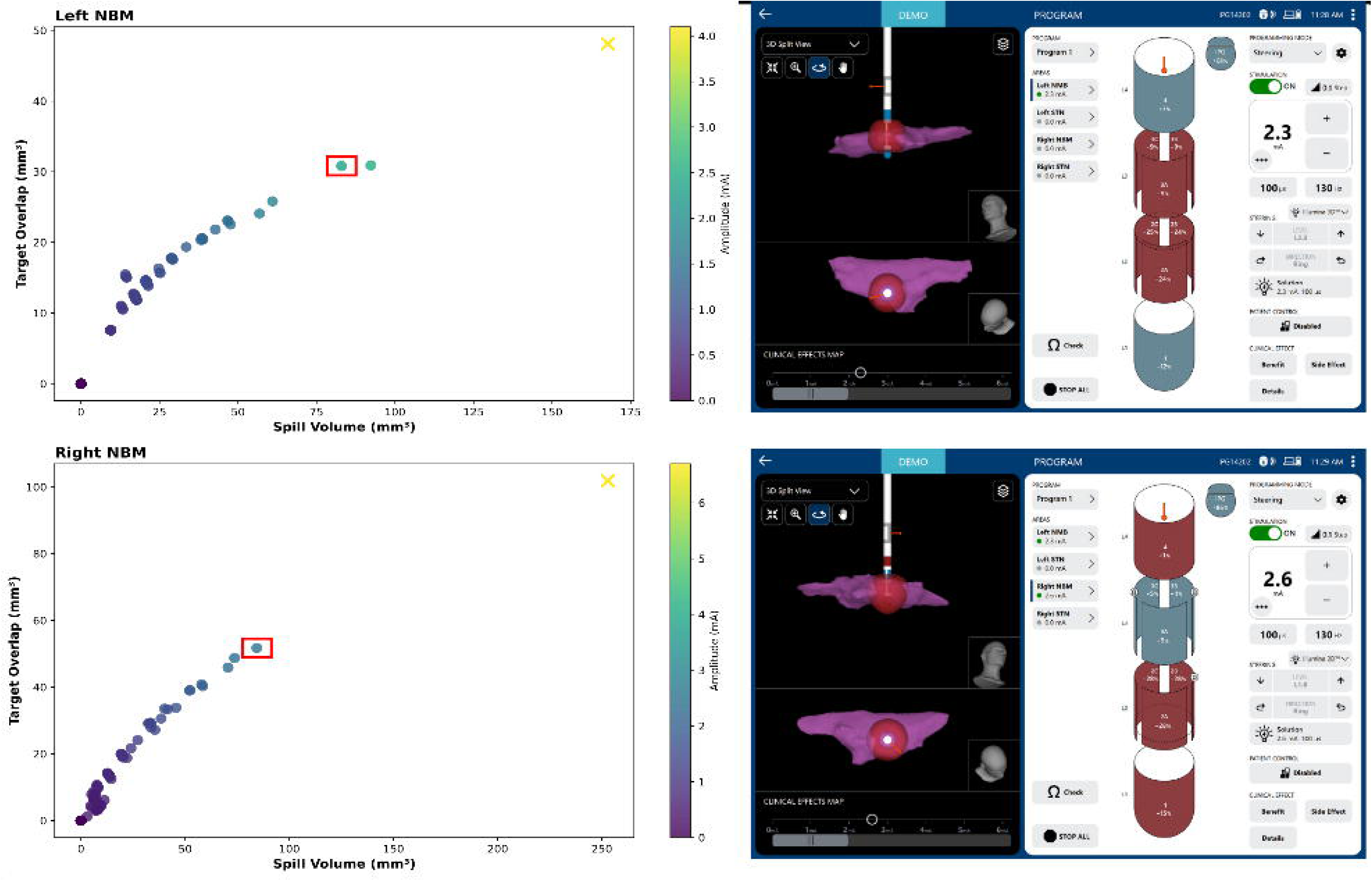
Image-Guided DBS Settings for the Vertical Trajectory. (Left) Depiction of all possible Illumina results from different weightings of avoidance and background ratios for the Left NBM (Top) and Right NBM (Bottom). Spill volume is shown on the x-axis and target (i.e., NBM) overlap is shown on the y-axis. The suggested stimulation amplitude is depicted by the color bar. Settings that exceeded the allowable overlap with ROAs are denoted as an ‘X’. The clinician-determined setting is shown by the red box. Only settings where Illumina converged on a solution are shown. (Right) Visualization of the DBS lead relative to the NBM (magenta) and the clinician-determined stimulation configuration and corresponding VTA (red).

**Figure 5.**
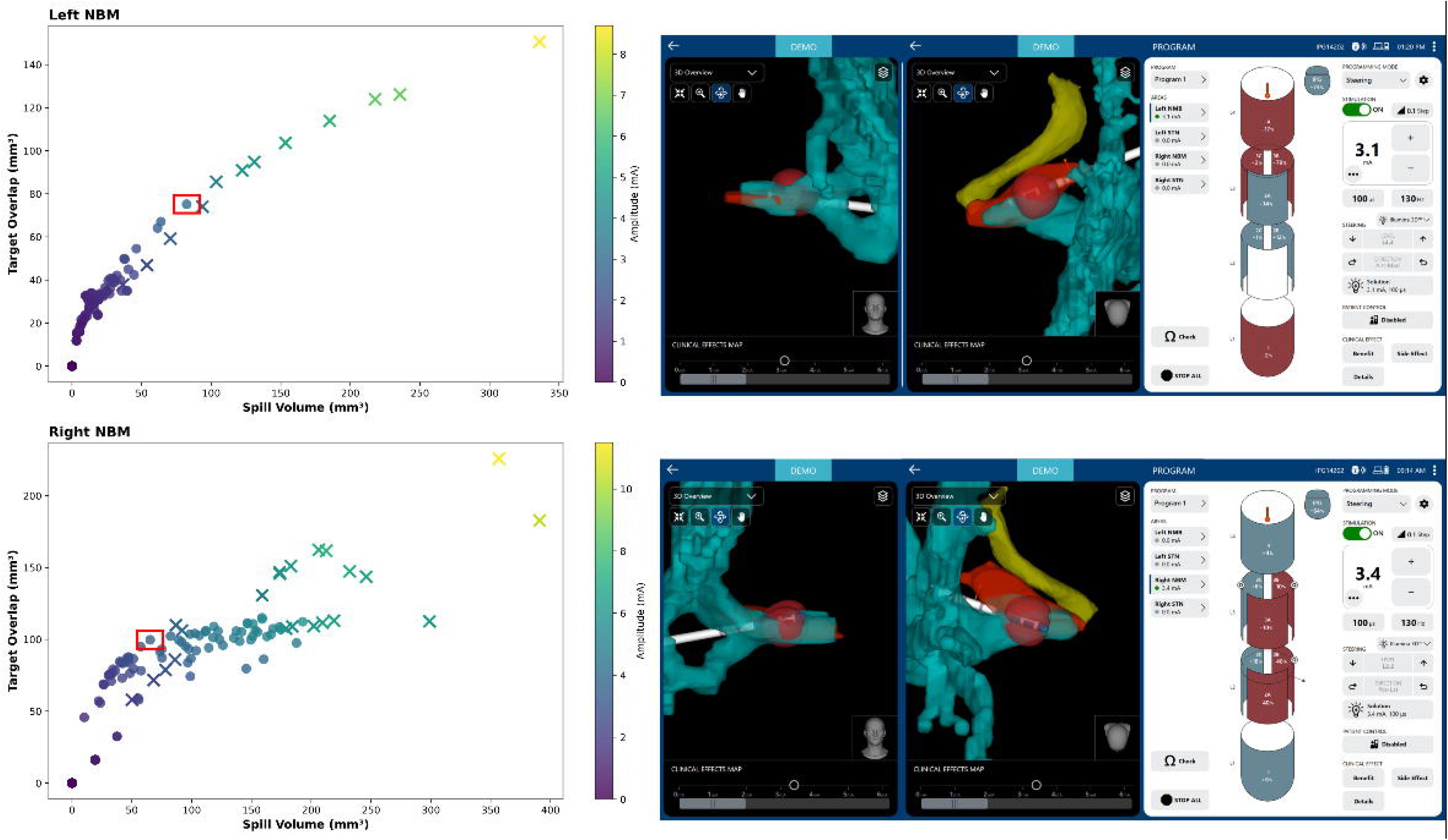
Image-Guided DBS Settings for the Lateral Trajectory. (Left) Depiction of all possible Illumina results from different weightings of avoidance and background ratios for the Left NBM (Top) and Right NBM (Bottom). Spill volume is shown on the x-axis and target (i.e., NBM and its lateral tract) overlap is shown on the y-axis. The suggested stimulation amplitude is depicted by the color bar. Settings that exceeded the allowable overlap with ROAs are denoted as an ‘X’. The clinician-determined setting is shown by the red box. Only settings where Illumina converged on a solution are shown. (Right) Visualization of the DBS lead relative to the NBM (red) and its lateral tract (cyan) and the clinician-determined stimulation configuration and corresponding VTA (red). Optic Tract shown in yellow.

### Safely Programmed without Adverse Events

The STN was successfully programmed at 130 Hz and 60 microsecond pulse width on Day one of the Initial Programming following standard clinical practice. On Day two, the NBM was programmed using an intermittent stimulation pattern of 60 Hz and 100 microsecond pulse width cycling 10 seconds ON and 40 seconds OFF and a symmetric biphasic active recharge pulse shape. The clinician tested the planned stimulation configuration identified by Illumina 3D™. Amplitude was increased in steps of 0.5 mA until the targeted amplitude was reached. No nausea, pulling or tingling, dizziness, imbalance, visual issues or phosphenes, cognitive symptoms, psychiatric symptoms, body temperature regulation issues or other symptoms were endorsed by any of the participants or clinician as part of separate customized symptom questionnaires. The NBM stimulation was imperceptible to all patients. After finalizing stimulation settings, the investigative Chronos software (Boston Scientific) was used to implement a schedule of 23 hours of STN only stimulation and 1 hour of combined STN and NBM stimulation with the NBM hour set to occur at approximately 10am. All participants were programmed successfully on the planned stimulations settings and sent home on both STN and NBM neurostimulation. No adverse events occurred during programming.

### Six-Month Outcomes

Outcomes were evaluated after six months of combined STN and NBM stimulation. All five participants improved from their off/OFF baseline MDS-UPDRS III done at Initial Programming compared to on/ON at 6 months (Average±SD at baseline compared to 6 months: 46.4±10.7 to 21.2±5.6).

The primary cognitive outcome was the Parkinson’s Disease - Cognitive Rating Scale (PD-CRS), which is a global neuropsychological evaluation designed to assess cortical and subcortical function. The PD-CRS includes nine subtests assessing sustained attention, working memory, immediate and delayed verbal memory, alternating verbal fluency, semantic verbal fluency, spontaneous clock drawing, figure-naming, and clock-copying tasks and is recommended by the International Parkinson and Movement Disorders Society (MDS) as a global cognitive assessment instrument for PD^55,56^.

At the baseline testing, done after the DBS procedure but prior to Initial Programming, three participants fell within the PD-CRS established PD-MCI (Parkinson’s Disease-Mild Cognitive Impairment) range, one participant fell within PD-D (dementia)^54^, and one fell within PD-normal. All five participants showed notable improvements in the PD-CRS at their six-month evaluation (Average±SD improvement: 12.6±4.0 points; Figure 6A). All 3 the PD-MCI range improved into the normal range, and the patient initially falling within the PD-D range improved into the PD-MCI range. All 5 participants exceeded the Minimally Clinically Important Difference of +7.7 points^57^ (Fig. 6B).

**Figure 6.**
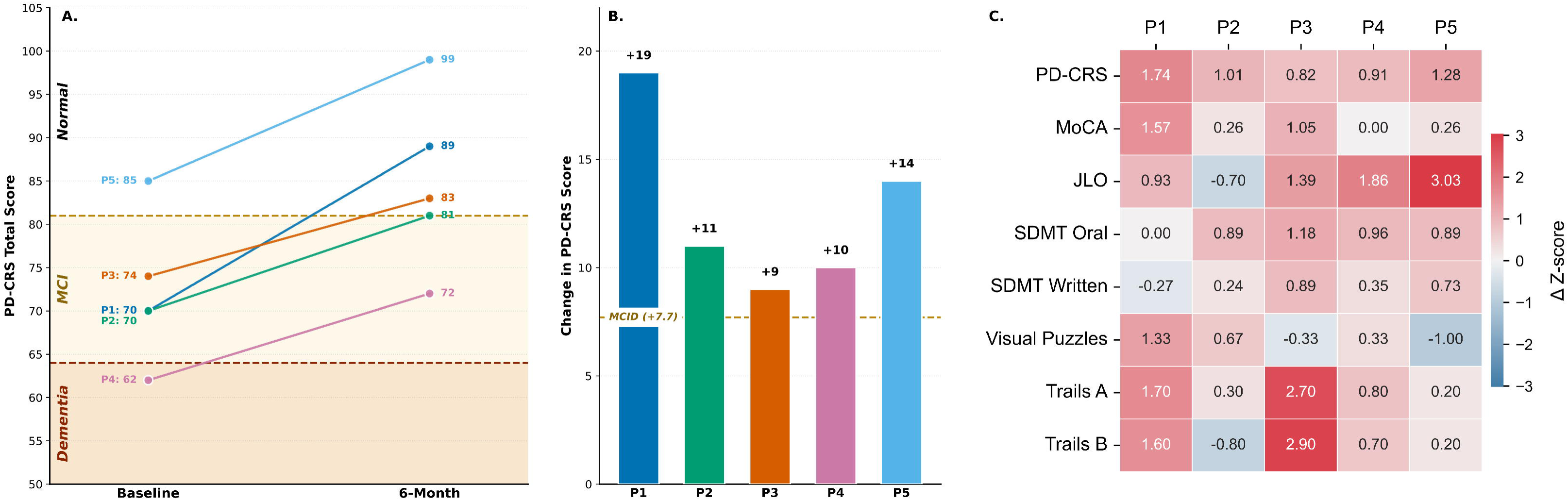
Six-Month Neuropsychological Outcomes. (A) Individual baseline and 6-month data for the Parkinson’s Disease – Cognitive Rating Scale (PD-CRS) with the PD-CRS established PD-Dementia and PD-MCI ranges highlighted. (B) Individual observed change in the PD-CRS relative to baseline in comparison to the established Minimally Clinically Important Difference. (C) Heatmap of the observed change in Z-score after 6 months across the neuropsychological assessments with positive (red) numbers indicating improvements.

Participants improved across nearly all neuropsychological assessments, with some variability among the group across domains (Fig. 6C). There was an overall improvement in all neuropsychological assessments, including the Montreal Cognitive Assessment (Average±SD Z- score change: 0.63±0.66), as well as in Executive function as measured by the Trails B (0.92±1.41), in Processing Speed/Attention as measured by the Symbol Digit Modalities Test Oral (0.78±0.45) and Written (0.39±0.45) and by Trails A (1.14±1.06), and in Visuospatial ability as measured by the Visual Puzzles from the Wechsler Adult Intelligence Scale-IV (WAIS-IV; 0.20±0.90) and by Judgement of Line Orientation (1.30±1.37). See Supplementary Table 2 for full results.

## Discussion

In this first in human study, we present the early results demonstrating clinically meaningful cognitive improvement after six months of intermittent patterned neurostimulation of the NBM in 5/5 participants with PD, PD-MCI and PD-D. The three participants with PD-MCI improved into the PD-normal range of the PD-CRS and the one PD-D participant moved into the PD-MCI range. All received bilateral STN and bilateral NBM DBS lead placement in a single procedure, with standard micro-electrode recording (MER) for the STN lead placement and image guided NBM target lead placement. A pre-clinical, evidence-based vertical trajectory for the NBM was used in three patients and a novel lateral trajectory targeting the immediate fiber outflow of the NBM lateral tract was used in two other patients. Intra- and post-operative lead reconstructions confirmed accurate lead placements relative to the STN and NBM targets.

Initial programming was performed following standard of care for STN DBS, which was on continuously at 130 Hz. Given the lack of behavioral symptom feedback to guide manual programming, an image-guided automated algorithm (Illumina 3D™) was used to determine the current and anodal or cathodal polarity on each NBM DBS electrode so as to maximize target overlap while minimizing overlap of nearby regions that may have elicited side effects. NBM DBS was on intermittently for one hour every morning. During that hour NBM DBS was ON (60Hz) for 10 seconds and OFF for 40 seconds. The stimulation programming was well tolerated by all patients.

Assessments following six months of combined continuous STN and intermittent NBM DBS showed substantial clinical improvements in all five participants, that exceeded the established Minimally Clinically Important Difference, Fig. 6B. The addition of the two leads in the NBM did not negatively affect the beneficial motoric effects of STN DBS as all five participants showed expected improvements with STN DBS in motor symptoms at 6-months compared to baseline (52.5±13.9% improvement in MDS-UPDRS III).

There are currently no FDA-approved treatments for PD-MCI. The only notable positive RCT for PD-D was for the cholinesterase inhibitor rivastigmine, which is currently only approved for mild-to-moderate PD-D and only demonstrated minor benefits (2.1±8.2 point or 8.8% improvement on the ADAS-cog), with only 19.6% of patients showing clinically meaningful improvements^6^. Furthermore, it was associated with bothersome side effects such as nausea, vomiting, and increased tremor. Rivastigmine failed to show efficacy in a single randomized placebo-controlled trial for PD-MCI^58^, as did the PD MAO-B inhibitor rasagiline^59^. Meanwhile, here we observed a mean improvement of 12.6±4.0 points on the global PD-CRS (17.5±5.7% improvement) with all 5 patients exceeding the established MCID after 6 months of combined STN and intermittent NBM DBS. This replicates previous findings in the non-human primate where intermittent NBM DBS significantly outperformed administration of the cholinesterase inhibitor donepezil^37^.

Previous attempts of NBM DBS for cognition have had variable results. The initial case study reported by Freund and colleagues in 2009 showed cognitive improvements in a PD-D patient in the Rey Auditory Verbal Learning Test and Trails A following a 4 lead STN and NBM DBS approach^43^. However, the patient remained in the severely cognitively impaired range. Meanwhile, two different randomized, double-blind crossover trials in 6 patients each with PD-D or PD-MCI showed no consistent improvements in cognitive outcomes using a combined 2-lead GPi and NBM DBS approach^44,47^. The major critical difference between these studies and the current study is the type of stimulation used for the NBM. The previous studies all used continuous low-frequency 20 Hz stimulation 24 hours per day. This was chosen based on the rationale that NBM neurons’ firing rate is typically around 20 Hz. However, evidence from the non-human primate and rodent both show that intermittent stimulation (60 Hz cycling ON for 10-20 second and OFF for ∼40 seconds) for only 1 hour a day improves cognitive performance, whereas continuous low frequency stimulation does not and can even impair performance^34,35,37,39^. The importance of this intermittent pattern of stimulation was recently demonstrated by Sasikumar et al., who found that 3 patients who previously received no benefit on continuous 20 Hz NBM DBS, showed improvements in attention on a computerized task after 4 weeks of intermittent NBM DBS^48^.

The temporal dynamics of cortical acetylcholine release suggests why intermittent cycling (60 Hz ON for 10-20 second and OFF for ∼40 seconds) for only 1 hour a day may be optimal for improving cognition. Continuous NBM stimulation leads to an initial peak of cortical acetylcholine within the first 20 seconds, but from there it begins to decline back to baseline or even lower within 3 minutes, likely due to depletion of acetylcholine reserves^38,52^. Meanwhile, an intermittent pattern such as 10-20 seconds ON followed by 40 seconds OFF enables the system to recapture the peak cortical acetylcholine response during each stimulation cycle^38^. Shanazz and colleagues investigated the dynamics of cortical acetylcholine release under different NBM stimulation settings in mice and found that 60 Hz and higher frequency stimulation led to significantly higher peak cortical acetylcholine release compared to 20 Hz; furthermore, 10 seconds of 60 Hz stimulation was sufficient to achieve peak acetylcholine release in the NBM, and that recovery occurred within approximately 15-20 seconds, which is likely governed by acetylcholine hydrolysis, diffusion, and choline transport^40^. Therefore, the intermittent stimulation pattern repeatedly perfuses the cortex with acetylcholine during each bout of stimulation followed by a recovery period.

Only one hour of intermittent NBM DBS, carried out automatically by the Chronos software, was used in the current protocol, as opposed to the continuous 20 Hz stimulation 24 hours/day used previously in the NBM DBS pilot studies. One hour has been sufficient to induce cognitive improvements in both the non-human primate and rodent models of dementia, whereas there was no further improvement seen with 24 hours of intermittent stimulation^34,35,37,39^. Given the key role the cholinergic system plays on sleep and arousal, constant stimulation may actually impair sleep, as it has been shown that NBM stimulation applied during sleep disrupts the normal sleep-wake cycle^60^. Additionally, periods of no stimulation may be necessary for the beneficial effect of increased nerve growth factor (NGF) and other neurotrophic factor release as increases in NGF release is seen 5 to 10 hours after NBM stimulation and is actually reduced during active stimulation periods, possibly due to increased NGF clearing from extracellular fluid due to the acute increase in cortical blood flow during stimulation^32^. It is also possible that the vasodilative effect is unable to be maintained for prolonged periods of time.

Several potential mechanisms may underlie the cognitive improvement observed following 6 months of intermittent NBM stimulation. NBM stimulation is expected to enhance activity and acetylcholine release in multiple cortical areas, such as the prefrontal, parietal, and temporal association cortices due to the diffuse cholinergic projections of the NBM. Animal studies have shown that the positive cognitive effects are cholinergic-dependent^35^. It also has a well-established vasodilative effect due to cholinergic receptors on blood vessels at NBM projection sites, which is important since increased cerebral blood flow is thought to be neuroprotective as it can prevent cell death and also supports clearance of potentially toxic metabolites^30,31,61^. Stimulation of the NBM may act not only on cortical blood vessels to increase cortical blood flow, but also on cortical neurons to enhance the release of neurotrophic factors, such as NGF, which acts as an endogenous neuroprotective factor in the cerebral cortex^32^. NGF prevents glucose deprivation-induced or glutamate-induced neuronal damage in cortical cell cultures^62,63^. Cholinergic neurons in the NBM itself are also dependent on extracellular NGF for maintaining their properties, and thus stimulation of the NBM may be beneficial both at the cortical and subcortical target regions of NBM projection axons, but also the NBM itself^64,65^. NBM DBS in the rodent has even been shown to increase cholinergic fiber length in the cingulate cortex and neurogenesis in the granule layer of the dentate gyrus^36^.

A major novelty of the current approach was the creation of an optimal trajectory for targeting the NBM that considered its unique geometry and anatomy. All studies to date in both humans and animals have utilized some variation of a vertical trajectory such that the axis of the DBS lead is almost perpendicular to the NBM. From the perspective of maximizing the VTA with the NBM, this trajectory is not ideal, as the NBM is inherently thin in the dorsal/ventral plane (only 1-3 mm) and spread approximately 13-14 mm antero-posteriorly and 16-mm medio-laterally^9^. Even with directional electrodes and steered current, there is no way to overlap a significant portion of the NBM with a vertical approach without significant overlap of neighboring regions such as the globus pallidus, putamen, amygdala, and/or optic tract. Therefore, we set out to design a trajectory that was optimal for the NBM anatomy given current DBS hardware constraints. Since the NBM funnels its projection fibers via 2 major fiber bundles: the medial and lateral tracts, this provided a mechanism to have widespread impact on the cortical cholinergic system using a directional electric field or VTA that encompassed most of the lateral efferent bundle as it exited the NBM^10^. We specifically targeted the lateral tract rather than the medial tract as there is greater evidence for its degeneration in PD, it projects to more cognitively relevant cortical areas and is easier to access surgically given its lateral location^20,50^. Tractography-based DBS approaches have grown increasingly popular as it is a means to have a large network effect with a small electric field due to the tight bundling of axons prior to their radiation to cortical areas^66–69^. Given the thin nature of the NBM, rather than approach the lateral tract vertically, we opted to approach from a lateral angle to align the axis of the DBS lead with the outflow tract and maximize overlap between the VTA and the immediate outflow of the lateral NBM tract in a subset of participants. This approach was then modified for practical surgical considerations. Specifically, the entry point was shifted to just posterior to the ear, creating a slight posterior-anterior angle. This served three purposes: first, a straight on trajectory would have necessitated an entry point from in front of the ear near the temple which was not practical; second, it allowed for coverage across the maximal dimension of the NBM lateral tract, thus theoretically maximizing the number of fibers modulated; and third, this avoided a trajectory through the insula and inferior limiting sulcus. Additionally, the dorsal-ventral trajectory was optimized to ensure the middle cerebral artery was avoided. The directional electrodes allowed us to prioritize steering current dorsal towards the NBM and its lateral tract and away from the amygdala, which is placed just ventral to the NBM.

## Limitations

The current study used an open label design in a small sample as it is a pilot safety and tolerability study which may lead to a placebo effect. A six-month interval and alternate forms were used between neuropsychological assessments to minimize practice effects and the primary neuropsychological assessment (PD-CRS) was not done as part of the evaluation neuropsychological battery. An alternative possibility is that the improvement seen at 6-months is a return to baseline after an initial deterioration from the surgery. Neuropsychological testing was done approximately one month after surgery to try and minimize this possibility and no clinical worsening in cognition was noted. Any image-guided approach is susceptible to potential registration errors or misidentified electrode orientations. We minimized potential errors due to brain shift by collecting a secondary CT scan approximately one month after surgery. However, detecting lead orientation for the lateral trajectory had greater uncertainty since the trajectory falls closer in plane to the CT gantry. The Loop-X® scanner (Brainlab) allowed us to minimize this issue as it could be rotated to a more orthogonal position to the trajectory, but this scan was done intra-op and immediately following the surgery. Additionally, the Illumina 3D algorithm does not incorporate fiber activation as part of its stimulation field modeling, nor does it consider frequency.

## Conclusion

This first-in-human study of intermittent neurostimulation of the NBM and its outflow resulted in improvement in cognition in five people with Parkinson’s disease. All participants showed clinically meaningful improvements in cognition after six months of combined STN and NBM DBS such that they moved from the PD-MCI to PD-normal range of the PD-CRS, or from the PD-D to PD-MCI range. Both a pre-clinical based vertical trajectory targeting the anterior portion of the NBM and a novel lateral trajectory targeting the immediate outflow of the lateral NBM tract were successfully implemented. Stimulation parameters from an automated imaging-based pipeline based on lead position and patient-specific anatomy was well tolerated in all participants. These results are preliminary but impactful in that they present the first evidence of reversal of cognitive decline in people with Parkinson’s disease and suggest that this procedure can be a safe adjunct to STN DBS for motor impairment in PD and should be performed prior to the development of PD-D.

## Methods

### Human Subjects

Five participants (3 female and 2 males) with clinically established Parkinson’s disease were enrolled in the study as part of a pilot clinical trial for combined STN and NBM DBS in PD (NCT05968703). Inclusion criteria included a) has a diagnosis of clinically established PD, b) been approved for STN DBS, c) >30% improvement in MDS-UPDRS III between off and on meds, d) Hoehn and Yahr stage <5, e) between 21-80 years old, f) presents with impairment in at least one test from comprehensive neuropsychological testing (i.e., performance at least 1 to 2 standard deviations below age, education, gender, and culturally appropriate norms, significant decline on serial cognitive testing, or significant decline from estimated premorbid levels), g) shows deficits in cholinergic aspects of gait (i.e., log swing time coefficient of variation ≥ 2.25 off therapy during stepping in place task^25^). Exclusion criteria included a) a diagnosis of dementia, b) unstable medical or psychiatric conditions, c) a history of seizures, d) untreated psychiatric conditions, e) a neurological or orthopedic disease that interferes with the ability to perform study tasks, f) implanted neurostimulators and MR incompatible metallic implants, g) drug or alcohol abuse. Use of Acetylcholinesterase inhibitors was not an exclusion criterion, but if a participant was actively taking them prior to enrollment, they were required to maintain the same dosage for the duration of the trial. Participants gave written consent to participate in the study, which was approved by the Food and Drug Administration with an Investigational Device Exemption and by the Stanford University Institutional Review Board.

### Study Design

The current study is part of a single-site open label pilot clinical trial testing the safety and tolerability of the combination of continuous STN DBS and intermittent NBM DBS for cognitive and cognitive-motor deficits in people with Parkinson’s disease. The study will involve 10 participants in total. Half the participants will receive a ‘Vertical’ trajectory targeting the anterior portion of the NBM using the same burr hole as the STN, and the other half will receive a ‘Lateral’ trajectory targeting the lateral efferent NBM fiber bundle from an entry point behind the ear. The current study reports on the feasibility of each of these surgical approaches in the first five participants. Interim analysis of neuropsychological outcomes was required at 6 months as part of the study protocol to ensure no significant cognitive deterioration occurred on NBM DBS that would require turning off the NBM leads.

#### Study Pipeline

All potential participants are first approved for STN DBS through the standard multidisciplinary Stanford DBS review board. Participants undergo standard clinical preoperative imaging for target identification and surgical planning. The participants receiving the Lateral trajectory also underwent a research MRI at the Stanford Lucas Center for Imaging. The anterior NBM target was identified on the structural imaging, and the lateral efferent fiber bundle was reconstructed using probabilistic tractography from the diffusion imaging. Participants underwent a modified two stage DBS implantation procedure with all 4 leads placed in stage one, followed by the implantable pulse generator in stage two. Lead localization was confirmed via in-surgery imaging, immediate post-op CT scans, and a follow-up 1 month CT scan. The lead location and patient-specific imaging was incorporated into Illumina 3D to determine the optimal stimulation parameters for the NBM. These stimulation parameters were tested at the Initial Programming approximately one month after the DBS procedure.

#### Vertical Trajectory

A vertical trajectory was utilized to target the anterior portion of the NBM. These leads use the same burr hole as the STN leads. This trajectory was modified from previous literature in the non-human primate that demonstrated consistent cognitive improvement when targeting 2 mm posterior to the crossing of the globus pallidus by the anterior commissure, and in the floor of the globus pallidus at the intersection of the GPi and GPe. The current target followed suit, while considering human histological evidence for the highest density of cholinergic neurons approximately 4-5 mm posterior to the anterior commissure.

#### Lateral Trajectory

Vertical NBM trajectories have been used in previous approaches but may not be optimal for targeting the NBM. Considering the area is a horizontal, diffuse, nearly flat area of the basal forebrain, a lateral approach would better maximize the potential surface contact with the lead and overlap with the subsequent produced electric field. In addition, the lateral efferent bundle, which projects to vast portions of the cortex, is located on the lateral side of the NBM, and thus a readily available target. Therefore, a lateral trajectory would allow for increased stimulation of both the NBM and its lateral efferent bundles, thus maximizing widespread cortical acetylcholine release.

### Preoperative MRI Acquisition

Multiplanar and multisequence preoperative clinical MRIs were acquired approximately one month before the surgery before and after intravenous administration of gadolinium-based contrast agent. MRIs were conducted on a 3T Discovery MR750 with an 8-channel head coil or SIGNA Premier with a 48-channel head coil (GE Healthcare) at the Stanford Neuroscience Health Center. Scan parameters can be found in Supplementary Table 1. The participants receiving the lateral trajectory underwent an addition research MRI. Diffusion-weighted images were acquired on a 3T GE SIGNA Premier scanner (GE Healthcare) with a 48-channel head coil at the Lucas Center for Imaging. A single-shot spin-echo echo-planar imaging (SE-EPI) sequence was employed with the following parameters: 60 diffusion-encoding directions at b = 1,000 s/mm^2^ and 6 interleaved b = 0 s/mm^2^, TR/TE = 7200/55.4 ms, flip angle = 90°, FOV = 220x220 mm^2^, acquisition matrix = 110x110, isotropic voxel size = 2.0x2.0x2.0 mm^3^, no interslice gap. An additional short reverse phase-encoded (posterior-anterior) b = 0 acquisition was collected for susceptibility-induced EPI distortion correction.

### Target Identification

A combination of T1-weighted, T2-weighted, and fGATIR images were used for identification of the NBM and surrounding landmarks (anterior commissure, optic tract, hippocampus, globus pallidus internus, globus pallidus externus, pallidum, etc.). The target for the Vertical trajectory was approximately 4 mm posterior to anterior commissure, laterally at the medial medullary lamina separating the GPe and GPi, and just ventral to the floor of the pallidum. For the lateral trajectory, the DWI research scan was used to implement probabilistic tractography to reconstruct the lateral NBM tracts, which as then integrated into the Brainlab neuronavigation software. A detailed explanation of the tractography method can be found in Crocket et al., 2023^50^. The target was the intersection between the NBM and the immediate outflow of the efferent fiber bundle where the entry point starts in the inferior temporal gyrus and approaches the lateral efferent bundle above the amygdala and piriform cortex.

### Surgical Procedure

All four leads for the bilateral STNs and NBMs were placed in one procedure for both the vertical and lateral trajectory. The 8-contact Boston Scientific Vercise™ Cartesia™ DBS leads (DB-2202) were used, as well as the Vercise Genus™ R32 IPG. Below are the specific details for each trajectory.

#### Stage 1: NBM Vertical Trajectory

An initial intraoperative CT scan was obtained after placement of 5 bone fiducials, which was then loaded into the StealthStation™ (Medtronic Inc.) intraoperative workstation and image fusion was performed between this, the preoperative CT scan, and the multiple MRI scans. A target was chosen in the STN bilaterally at approximately 11 mm lateral to the ACPC line, 5 mm inferior to the ACPC plane, and 3 to 4 mm posterior to the midpoint of ACPC. Targets were also chosen in the NBM bilaterally at approximately 18 mm lateral to the ACPC line, 6-7 mm inferior to the ACPC plane, and 10 mm anterior to the midpoint of ACPC. A reference arc was then attached to the FESS frame, and the entry points were located. A 1 mm twist drill was used to make small divots in the skull and then 14 mm burr holes were drilled bilaterally, and 2 SureTek bases were screwed into place. The bases of both Nexframe towers were screwed into the skull, and a reference arc was attached to the left and right Nexframe tower. The trajectory was aligned to each STN target and the guide cannula was lowered. MER was conducted according to standard practices, and an O-arm scan was done to confirm placement. The DBS electrodes were placed and the outer cannula withdrawn. Following securing of the STN leads, the tops of both Nexframes were reattached and the trajectory was aligned to the left and right NBM target. The guide cannula was lowered, and an O-arm scan was done to confirm placement, followed by introducing the two DBS electrodes. The DBS electrodes were placed and the outer cannula withdrawn. The leads were placed into their slots in each SureTek base and caps applied.

#### Stage 1: NBM Lateral Trajectory

An initial intraoperative CT scan was obtained after placement of 5 bone fiducials, which was then loaded into the ROSA ONE® robot (Zimmer Biomet) and image fusion was performed between this, the preoperative CT scan, and the multiple MRI scans. Image-guided computer-assisted targeting and trajectory were confirmed. The patient was placed supine in the Leksell head holder and registered the robot using the bone fiducials and confirmed sub millimetric registration accuracy. Subthalamic nucleus leads were planned and executed according to standard of care. For the NBM trajectories, two additional small linear incisions were planned and twist craniostomy holes were made using Rosa guidance. The left NBM was the first target. The guide cannula was lowered to 15 mm above the final target, and an initial Loop-X spin was taken to confirm accuracy. The 8-contact lead was measured to the appropriate length and placed to target, and the outer cannula was withdrawn. The lead was fastened with a silastic sleeve and dog bone anchor. This process was repeated for the right NBM trajectory, and another Loop-X spin was taken to confirm accuracy of both leads. The STN trajectories were carried out after the NBM and included MER and standard processes using the Rosa robot. A final Loop-X spin was conducted to confirm accuracy of the four leads.

#### Stage 2 Procedure

For stage 2, the patients were brought to the operating room where general anesthesia was induced. They were then placed into the supine position with a roll under the left shoulder. The head was turned toward the right. The chest and left side of the neck and scalp were shaved, prepped and draped in the usual sterile fashion. After infiltration of a half-and-half mixture of 0.5% Marcaine and 1% lidocaine with 1:100,000 epinephrine, a linear incision was made over the connectors. The DBS electrodes were located and the connectors were externalized, the set screws were loosened, and the connectors were removed from the leads and discarded. At this point, an incision was planned approximately 2-1/2 fingerbreadths below the left clavicle, and this area was infiltrated with local anesthetic solution. Following skin incision, a pocket was made in the plane above the pectoralis fascia for the pulse generator. The pulse generator was tested and was found to fit well within the pocket. Meticulous hemostasis was achieved with bipolar cautery. A subcutaneous pass was made from the scalp incision down to the chest incision, and 2 dual 60 cm extensions were brought through. The leads were attached to the proximal ends of the extensions, and the set screws tightened with a torque wrench. The distal ends of the extensions were inserted into the pulse generator, and the set screws tightened. Channels L1 and R1 were the left and right NBM leads, and channels L2 and R2 were the STN leads. The pulse generator was then placed into a TYRX antibacterial envelope. The excess extension was coiled beneath the pulse generator which was placed into its pocket. Impedances were tested and were found to be in an acceptable range. Both incisions were then irrigated copiously with bacitracin solution and closed in multiple layers using inverted interrupted sutures of 3-0 Vicryl for the subcutaneous tissue, inverted interrupted sutures of 2-0 Vicryl for the galea, staples for the scalp incisions and 4-0 nylon for the chest wound. It should be noted that prior to closure, vancomycin powder was sprinkled into the scalp incision. The wounds were dressed in a sterile fashion. The patient was awakened from general anesthesia and brought to the post anesthesia recovery room awake, alert, and unchanged from their preoperative neurological condition.

### Postoperative CT Acquisition

Following completion of the surgery, an immediate post-operative head CT scan was performed following the standard clinical protocol at Stanford. Scans were conducted on a Revolution CT scanner (GE Healthcare) in axial mode with a slice thickness of 0.625 mm, spacing between slices of 0.625 mm, FOV of 248 mm, matrix size of 512x512 mm and Tube voltage of 120 kVp. A secondary post-operative CT scan was performed approximately one-month post-surgery with the same settings to account for potential lead movement due to brain shift.

### Lead Localization

Leads were localized using Brainlab Elements Image fusion module (Brainlab, Munich, Germany) lead localization software, combining both the pre-operative MRI and post-operative CT scans. The ideal CT scan for performing lead localization should use slices as orthogonal to the leads as possible. The orientation of the lateral trajectory means the leads are closer to parallel with the axial slices on a typical head CT scan, leading to suboptimal artifact generation for determining lead position and orientation. To remedy this issue, CT scans completed during surgery were used to aid determination of lead position and orientation. The Brainlab Loop-X allowed for rotation of the gantry to the desired orientation, improving resolution of the lead position. Lead position and orientation was confirmed by an expert rater (VC).

### DBS Illumina 3D Algorithm

The DBS Illumina 3D™ algorithm aims to optimize the stimulation parameters by considering user-set regions of interest (ROI) and regions of avoidance (ROA), maximizing ROI stimulation and minimizing ROA stimulation. Choice of ROA/s can be advised by characteristics unique to the ROI and the surrounding areas where stimulation could cause adverse side effects. The Illumina 3D™ algorithm incorporates the Boston Scientific directional DB-2202 Cartesia leads. For each DBS lead, the optimal amplitude and fractionalization (i.e., the unique arrangement of current driven to each electrode, expressed as a percentage of the total current) are determined from the user-set weighted importance of the fill of the ROI stimulation relative to stimulation spill, as well as the penalty for overlap with ROAs. Additional details regarding the Illumina 3D™ algorithm can be found in Malekmohammadi et al., 2022^53^. For the vertical trajectory, the NBM was set as the ROI and for the lateral trajectory, a combined single object of the NBM and the lateral tract was set as the ROI. The ROAs were set to Optic Tract, Amygdala, and Globus Pallidus Externus due to their proximity to the NBM and potential for eliciting side effects.

### Selection of Stimulation Parameters

The Illumina 3D™ algorithm was run across 11 different weightings for both fill/spill and ROI/ROA ratios, resulting in a total of 121 different potential stimulation configurations. For each configuration, the percent overlap with the ROI and each ROA was determined, as well as the overall spill (i.e., portion of the stimulation field that does not overlap with the ROI). Stimulation configurations with excessive overlap of ROAs were not considered. Visualizations were created of the remaining potential configurations comparing the target overlap compared to overall spill volume to identify the best candidates that efficiently overlapped with the target. Visualizations of the resulting Stimulation Field Model (SFMs) for the candidate configurations alongside the nearby ROAs were then produced. The clinician (HBS) determined the final optimum stimulation configuration considering maximization of overlap with the target, minimum overlap with nearby anatomy/ROAs, and overall efficiency of the SFM.

### Chronos

The Chronos Research Engine is an investigational software from Boston Scientific that allows complex control of patterned DBS in human subjects to a much finer level than permitted by commercial DBS systems. It allows multiple stimulation programs to be scheduled to turn on and off at pre-specified times. Chronos works with existing Boston Scientific commercial Vercise™ Genus™ rechargeable pulse generators and connected DBS leads, and patients can switch from investigational to commercial DBS programs with their regular patient programmer. No change (no new firmware) is required for the stimulator: Chronos simply permits access to stimulation capability that already exists in the Vercise Genus stimulators. All stimulation delivered with Chronos obeys commercial safety limits on charge density, total charge, and amplitude. Chronos was used to deliver the scheduled intermittent stimulation necessary for the current paradigm. The use of Chronos was FDA IDE approved.

### Initial Programming

Initial Programming occurred approximately one month after the DBS surgery. It was split across two days. The first day followed standard clinical practice for programming the STN off medication. The NBM programming was performed on the second day with STN DBS turned ON and on medication. Stimulation of the NBM was not expected to produce immediate symptoms to determine the effectiveness of the stimulation. Therefore, during the programming only possible side effects were tested such as the stimulation of the optic tract, nausea, mood changes, body temperature regulation issues, tingling, pulling, dizziness, or tightness of the muscle of the face or arms. Programming was performed with Boston Scientific’s investigational Chronos software. This software allows for different programs to be scheduled for a certain amount of time. For programming, Program 2 contained only the STN leads and Program 3 contained both the STN and NBM leads. Program 2 was set to run for 23 hours and program 3 was set to run for 1 hour, allowing for an automatic switch on and off for the NBM leads for 1 hour per day. Chronos also allows cycling to be used which is required for the NBM stimulation. Cycling was set to 10 seconds ON and 40 seconds OFF with a 100-microsecond pulse width. A symmetric biphasic pulse (i.e., active recharge) was used. Left and right sides were turned on separately during programming with 0.5 mA increments for each side until the targeted amplitude was reached. After each increase, the participants were asked if they experienced any of the abovementioned symptoms. If they did not, then a test was performed to check eyesight and motor symptoms. Stimulation was subsequently increased and tested again until reaching the final selected stimulation amplitude. Once both hemispheres were set, the participant sat resting for fifteen minutes to allow for stimulation wash-in and to see if any adverse events occurred from the stimulation of the NBM.

### Neuropsychological Assessments

Neuropsychological assessments were conducted approximately one month following the DBS surgery but before Initial Programming, as well as after six months of STN and NBM DBS. Testing was done in the on-medication state. For the 6-month assessment, STN DBS was also ON. The assessment battery included the Parkinson’s Disease – Cognitive Rating Scale (PD-CRS), Montreal Cognitive Assessment (MoCA), Judgement of Line Orientation (JLO), Symbol Digit Modalities Test (SDMT) both oral and written, Visual Puzzles from Wechsler Adult Intelligence Scale-IV, and Trails A and B.

## Data Availability

De-identified data from the clinical trial will be made openly available on the Data Archive for the BRAIN Initiative (DABI) within one year following completion of the trial: 10.18120/mm4v-s963.

## Acknowledgements

We thank Jeff Solomon and Wendy Gu for their technical expertise and support, Emilia Lambert, Jillian Melbourne, and Pranav Akella, as well as the participants who dedicated their time to this study.

## Author Contributions

Conceptualization: K.B.W, M.Z., D.T.B., J.M.H., V.P.B, H.B.S; Methods: K.B.W., P.A., C.C., R.A.C., G.K.D., M.M., H.B., A.G.R., M.Z., J.M.H., V.P.B; Software: K.B.W., R.A.C., M.M., H.B.; Validation: G.K.D.; Formal Analysis: K.B.W., P.A., C.C., A.K.A., M.M., V.C., B.R.; Investigation: P.A., C.C., A.K.A., G.S., T.M.H., M.P.B.; Resources: H.B., M.M., B.R.; Data Curation: P.A., C.C., A.K.A., M.P.B.; Writing – Original Draft: K.B.W., P.A., C.C., H.B.S.; Visualization: K.B.W., C.C.; Supervision: K.B.W., H.B.S; Project Administration: K.B.W., P.A., C.C., A.K.A., M.P.B.; Funding Acquisition: H.B.S. All authors reviewed, commented, and approved the manuscript.

