## Supplementary Materials for "Clinically Meaningful Cognitive Improvement from Intermittent Neurostimulation of the Nucleus Basalis of Meynert in Parkinson’s Disease"

**Supplementary Table 1.** MRI Sequence Parameters

| Parameter | T1 BRAVO | T2 CUBE | fGATIR |
| --- | --- | --- | --- |
| TR (ms) | [7.284 – 7.608] | 2500 | [7.284 – 7.612] |
| TE (ms) | [2.996 – 3.16] | [85.31 – 85.962] | [2.996 – 3.16] |
| TI (ms) | [400 – 600] | - | 450 |
| Flip angle (°) | [9.0 – 13.0] | 90.0 | 8.0 |
| FOV (mm) | 240 | 240 | 240 |
| Matrix | 256x256 | 320x320 | 256x256 |
| Slice thickness (mm) | [1.0 – 1.12] | 1.0 | 1.0 |
| Space Between slices | 1 | 0.5 | 0.5 |
| Voxel size (mm^3^) | 0.47x0.47x [1.0 – 1.12] | 0.47x0.47x1.0 | 0.94x0.94x1.0 |
| Acquisition plane | Axial | Sagittal | Axial |

**Supplementary Table 2.** Neuropsychological Assessments

| Assessment | P1 | | P2 | | P3 | | P4 | | P5 | |
| --- | --- | --- | --- | --- | --- | --- | --- | --- | --- | --- |
|  | Baseline | 6-Month | Baseline | 6-Month | Baseline | 6-Month | Baseline | 6-Month | Baseline | 6-Month |
| PD-CRS | 70 | 89 | 70 | 81 | 74 | 83 | 62 | 72 | 85 | 99 |
| MoCA | 17 | 23 | 22 | 23 | 18 | 22 | 14 | 14 | 24 | 25 |
| JLO  (Z-score) | -0.77 | 0.16 | -1.24 | -1.94 | -1.47 | -0.077 | -4.27 | -2.57 | -3.80 | -2.41 |
| SDMT – Oral  (Z-score) | -1.32 | -1.32 | -2.96 | -2.07 | -2.06 | -0.88 | -2.87 | -2.41 | -1.17 | -1.91 |
| SDMT – Written (Z-score) | -0.67 | -0.94 | -2.12 | -1.88 | -2.00 | -1.11 | -2.53 | -1.91 | -1.76 | -2.18 |
| Visual Puzzles (Scaled score) | 9 | 13 | 7 | 9 | 7 | 6 | 6 | 7 | 7 | 4 |
| Trails A  (T-score) | 38 | 55 | 39 | 42 | 24 | 51 | 39 | 47 | 47 | 49 |
| Trails B  (T-score) | 38 | 54 | 49 | 41 | 19 | 48 | 14 | 21 | 43 | 45 |

PD-CRS: Parkinson’s Disease – Cognitive Rating Scale; MoCA: Montreal Cognitive Assessment; JLO: Judgement of Line Orientation; SDMT: Symbol Digit Modalities Test

**Supplementary Table 3.** Individual NBM DBS Settings

| ID | Side | Amp  (mA) | E1 | E2 | E3 | E4 | E5 | E6 | E7 | E8 |
| --- | --- | --- | --- | --- | --- | --- | --- | --- | --- | --- |
| P1 | L | 2.3 | 12 | -24 | -24 | -25 | -9 | -9 | -9 | 7 |
|  | R | 2.6 | -15 | -28 | -28 | -28 | 5 | 4 | 5 | -1 |
| P2 | L | 3.1 | -2 | - | 12 | 1 | 14 | -79 | -2 | -17 |
|  | R | 3.4 | 8 | -40 | -40 | 18 | -10 | -10 | 6 | 4 |
| P3 | L | 3.6 | -2 | 1 | - | 11 | -4 | 14 | -82 | -12 |
|  | R | 3.1 | 8 | -4 | -85 | 9 | -1 | -10 | 2 | 3 |
| P4 | L | 2.3 | 8 | -31 | -31 | -32 | -2 | -2 | -2 | 3 |
|  | R | 2.3 | 4 | -46 | -50 | 23 | -2 | -2 | 3 | 2 |
| P5 | L | 3.2 | 9 | 15 | -23 | -52 | 6 | -8 | -17 | 6 |
|  | R | 3.8 | -7 | -10 | -82 | 16 | 1 | 9 | 0 | -1 |

* All 60 Hz and 100 μs pulse width, cycling 10 seconds ON, 40 seconds OFF for 1 hr/day. E1-8 refer to the individual contacts on the 8-contact lead.
